# The risk prediction models of neonatal necrotizing enterocolitis in China: a systematic review

**DOI:** 10.1101/2024.02.04.24302320

**Authors:** Jiaming Wu, jiajia Du, Junjie Peng, Xin Guo, Xue Hu, Yunchuan Li, Yuanfang Wu

## Abstract

**Objective:** Systematically evaluate the risk prediction model for neonatal necrotizing enterocolitis (NEC) in China, providing reference for clinical work and future research.

**Methods:** We searched Chinese and English databases were systematically searched to focus on NEC risk prediction modeling studies.The search time ranged from database establishment to 25 August 2023.Two researchers independently screened the literature and extracted information.Then risk of bias and applicability were assessed by using the Prediction Model Risk of Bias Assessment Tool.

**Results:** A total of 10 papers involving 12 NEC risk prediction models were included, which is focusing on the populations of preterm infants mostly, building the methods of models diversity, predicting factors discrepancy widely.

**conclusion:** The existing NEC risk prediction models in China have good predictive performance, while they often lack external validation, resulting in an overall high risk of bias. In the future, clinicians and nurses should learn from the evaluation criterions based on the PROBAST, then to test and verify them. Or machine learning algorithms usage is to construct models with operationalization and better predictive efficacy. [REGISTRATION: PROSPERO ID: CRD42024503844]

## 1. Introduction

NEC is one of the lethal diseases involving the gastrointestinal tract in newborns, which is mainly characterized by abdominal distension, vomiting, diarrhea, bloody stools, feeding difficulties, and in severe cases, shock, multi-organ failure, necrosis of the whole intestinal segment, and even death [1].According to data from the National Center for Health Statistics, NEC was one of the top ten causes of newborn mortality in the United States in 2019, with a mortality rate of 0.094‰, higher than the 0.079‰ in 2018[2].The report shows that the mortality rate of NEC in extremely low birth weight (ELBW) infants is as high as 30.0% to 50.9%[3].The pathogenesis of NEC is not yet clear, and its early clinical manifestations are relatively insidious. However, the disease progresses rapidly, with a high mortality rate and many sequelae. Even during the recovery period of NEC, recurrence is still a certain risk[4].

In addition, the lack of reliable biomarkers for NEC diagnosis [5] makes clinical diagnosis difficult and increases nursing risks. Therefore, early identification of high-risk children who may develop NEC and taking targeted preventive measures are of great significance for improving prognosis[6].In recent years, research on the development, validation, and application of NEC risk prediction models has been increasing both domestically and internationally. However, there are significant differences in the applicable population, predictive factors, actual application performance, and research methods of each research prediction model. This study provides a scope review of the construction, validation, predictive factors, and performance of current predictive models in China, which is aiming to provide reference for clinical nursing work and future research.

## 2. Method

This systematic review was conducted according to the Preferred Reporting Item (PRISMA) updated guidelines for systematic reviews and meta-analyses [6]. The protocol has been registered in the International Prospective Register of Systematic Reviews (PROSPERO) and the registration number is CRD42024503844.

### 2.1 Inclusion and exclusion criteria

Inclusion criteria:

1. The research subject is newborns.
2. The research focuses on developing or validating NEC risk prediction tools.
3. The research types include cross-sectional studies, longitudinal studies, cohort studies, case-control studies, etc.
4. Research is carried out in China.
5. Published Chinese or English literature publicly.

Exclusion criteria:

1. Duplicate publications.
2. Data on outcome indicators could not be extracted.
3. Review articles, case reports and letters to the editor.

### 2.2 Search strategy

Eight databases were chosen for a systematic search of studies, including PubMed, Embase, the Cochrane Library, Web of Science, CNKI, VIP, China Biology Medicine disc (CBM) and WanfangDatabase.The search time ranged from database establishment to 25 August 2023.The search was conducted using Medical Subject Headings (MeSH) and free terms,including infant *, premat-ure,neonatal prematurity, low birth weight infant, enterocolitis, necrotizing, necrotizing enterocolitis, risk prediction, predict*, risk assessment, risk factors, Nomograms, Nomogram, model*, tool*, scale*. Search terms were combined using boolean operators.Additionally, manual searches and citation searches were also done.Taking PubMed as an example, the search formula is as follows:

#1:”infant, premature”[MeSH Terms]

#2:”infant*”[Title/Abstract] OR “neonatal prematurity”[Title/Abstract] OR “lo w birth weight infant”[Title/Abstract]

#3:#1 OR #2

#4: “enterocolitis, necrotizing”[MeSH Terms]

#5: “enterocolitis necrotizing”[Title/Abstract] OR “necrotizing enterocolitis”[Title/Abstract]

#6: #4 OR #5

#7:”Nomograms”[MeSH Terms]

#8:”Nomograms”[Title/Abstract] OR “risk prediction”[Title/Abstract] OR “pr edict*”[Title/Abstract] OR “risk assessment”[Title/Abstract] OR “risk factors”[Title/ Abstract] OR “Nomogram”[Title/Abstract] OR “model*”[Title/Abstract] OR “tool*” [Title/Abstract] OR “scale*”[Title/Abstract]

#9:#7 OR #8

#10:#3 AND #6 AND #9

### 2.3 Literature screening and data extraction

After the search, the retrieved literature was imported into NoteExpress3.7 in order to remove duplicates. According to inclusion and exclusion criteria, two trained researchers read literature titles and abstracts for preliminary screening, and then we read intensively the full text again for the second sieve to choose.If there was disagreement,we will discuss and resolve the differences with the third researcher.This study developed a standardized information extraction table based on the Prediction Model Risk of Bias Assessment Tool (PROBAST) checklist to independently extract literature information[8]. The extraction content includes: first author, publication year, research object, study design, sample size, number of outcome events, missing data and processing methods, model building methods, discriminability (AUC/C-statistic/C-index) Calibration degree (calibration chart), model validation status, predictive factors, model presentation form, etc.

### 2.4 Risk of bias and applicability evaluation

Two researchers independently evaluated the methodology of the included literature according to the PROBAST checklist and cross checked it. PROBAST conducts bias risk assessment in four areas: research subjects, predictive factors, outcomes, and statistical analysis. The assessment results in each area have three levels: “low”, “high”and “unclear”, with a total of 20 signal problems.PROBAST conducts applicability evaluations in three areas: research subjects, predictive factors, and outcomes, using methods similar to bias risk assessment[8].

### 2.5 Statistical analysis

Descriptive analysis methods were used to organize and summarize the included literature, including the basic characteristics of the model, model establishment methods, model validation methods, and predictive performance. By extracting indicators such as AUC, sensitivity, and specificity to reflect the discrimination ability of the model (the ability to accurately distinguish between diseased and non diseased individuals), AUC value<0.7 indicates poor discrimination, 0.7 ≤ AUC value ≤ 0.8 indicates moderate discrimination, and AUC value>0.8 indicates good discrimination. By extracting calibration curves, Hosmer Lemeshow tests, and other indicators to reflect the calibration of the model (the accuracy of predicting the probability of future events occurring for an individual), it is used to evaluate the consistency between “predicted risk” and “actual risk”.

## 3. Results

### 3.1 Literature screening process and results

A total of 7948 relevant literature were retrieved,of which 3490 were duplicates.By reading thetitle and abstract, 4253 studies were eliminated and 205 studies were retained for full-text reading to assess eligibility. Ultimately, 10 studies were included.The literature screening process isdetailed in Figure 1.

**Figure 1.**
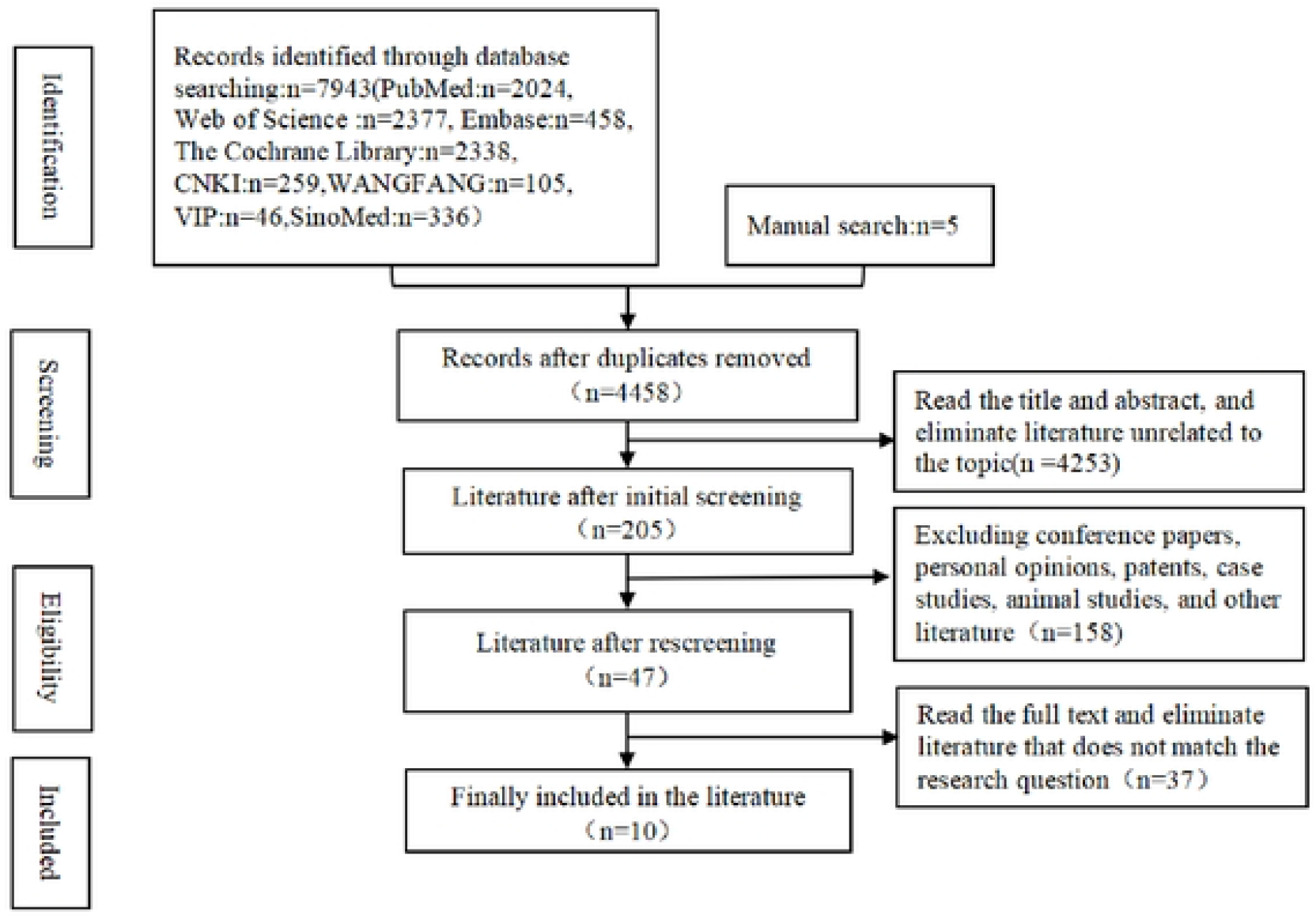
Flow chart of literature screening.

### 3.2 Characteristics of the included studies

The 10 included articles are all from China [9-18],with 7 retrospective cohort studies and 3 case-control studies.One of Wang’s [18]studies is a doctoral thesis, involving three predictive models, represented by “Wang 1”, “Wang 2”, and “Wang 3” respectively in the following text.The characteristics of the study are shown in Table 1.

### 3.3 Inclusion of literature risk of bias and applicability evaluation results

The 10 studies included have an overall high risk of deviation, and the applicability evaluation results show that the overall applicability is good.The results of bias risk assessment are shown in Table 2.

### 3.4 The construction method, predictive efficiency, and content of NEC risk prediction model

#### 3.4.1 Model establishment status

A total of 12 NEC risk prediction models were established in the 10 included literature, with a focus on premature infants and mostly single center studies,the total sample size is 177-1173.In terms of model building methods, ten models [9,11-16,18] only used logistic regression algorithm, one model [10] used four algorithms: logistic regression, support vector machine, multilayer perceptron, and extreme gradient boosting, and one model [17] used three algorithms: artificial neural network, decision tree, and extreme gradient boosting.The modeling is detailed in Table 3.

#### 3.4.2 The performance of the model

Five models [9-11,16-17] were validated internally only, four models [12-15] used both internaland external validation, and three models[18] did not report a validation methodology or did not perform validation.Nine models [9-10,13,15-18] reported that the area under the curve (AUC) of the working characteristics of the subjects ranged from 0.716 to 0.947, all of which were above 0.7. Three models [11-12,14] reported C indices ranging from 0.844 to 0.973, all above 0.8, indicating good predictive performance.Three models [10-11,17] did not report or evaluate model calibration, while the rest were evaluated and the model fit was good (P>0.05).The vastmajority of models report specific presentation forms, mostly presented in the form of nomogram.The performance evaluation of the model is detailed in Table 3.

#### 3.4.3 The predictive factors of the model

The final prediction model consists of 4-15 variables, and predictive factors with a frequency of ≥ 2 occurrences included: birth weight, gestational age, infection, artificial feeding, sepsis, congenital heart disease, breastfeeding speed, patent ductus arteriosus (PDA), hypoalbuminemia, anemia [10,12-16,18].Among them, prenatal use of dexamethasone, breastfeeding, probiotics, use ofpulmonary surfactant (PS), and platelet distribution width>11.8 fL are protective factors [10-11,16].Gao et al [17] were unable to obtain the relevant predictors in their study.The predictive factors of other models are detailed in Table 4.

## 4. Discussion

### 4.1 The overall risk of bias in existing research is high, and the quality of methodology needs to be improved

The research design, establishment methods, validation methods, statistical analysis, etc. of predictive models determine their predictive performance[19].According to the PROBAST evaluationcriteria [8], all models included in this study have a high risk of bias.The reasons for the analysis may be that the research data mostly comes from traditional case-control studies or retrospective cohort studies, inappropriate variable data processing, failure to avoid using univariate analysis to screen predictive factors, and events per variable (EVP)<10 for each variable, whichlead to a higher risk of bias.In addition, PROBAST is a bias risk assessment tool proposed byMoons et al [20] in 2019 for predictive model research. Most scholars may not have fully designed their research according to this assessment tool, which may also be one of the reasons for the high bias risk.Therefore, the following recommendations are made based on the PROBAST evaluation criteria:For research design, prospective cohorts or nested case-control studies should be used as much as possible;Try to avoid using single factor analysis methods to screen predictive factors as much as possible;In model development research, it is advisable to ensure that the EPV is ≥ 20 or the sample size in model validation research is ≥ 100 cases;Do notarbitrarily exclude variables with no statistical significance in univariate analysis;In terms of model presentation, the probability formula can be transformed into simplified scoring rules or online calculation tools that are easy to apply display forms.

### 4.2 There are significant differences in the predictive factors of the model, and the research on risk factors has been slightly improved

NEC is generally believed to be a gastrointestinal disease caused by multiple stimuli, which poses a significant threat to the safety of newborns, predictive factors can help improve clinical diagnosis and prognosis[22].In this study, it was found that the predictive factors of each model were not the same, and the most frequent risk factors were birth weight, gestational age, artificial feeding, sepsis, PDA, and anemia. Studies have shown that there is a negative correlation between newborn birth weight and gestational age and the incidence of NEC. The smaller the gestational age and weight, the poorer the gastrointestinal development and immune system of newborns, making them more susceptible to infection[18,21-22].The inflammatory response caused by infection is an important part of the pathogenesis of NEC, and sepsis is an acute systemic infection caused by pathogens, which can directly damage the small intestine mucosa or induce inflammatory reactions[23].The mechanism of action of PDA on NEC is not yet clear, which may be related to intestinal ischemia caused by the transfer of blood flow from abdominal organs to pulmonary circulation [24].When hemoglobin is ≤ 80 g/L, it can be diagnosed as severe anemia, decreased albumin can cause a decrease in plasma colloid osmotic pressure, leading to reduced multi organ perfusion and microcirculation dysfunction. In addition, albumin deficiency can also cause dysbiosis of gut microbiota, epithelial cell rupture, and impaired immune function [13,25-26].Artificial feeding of newborns is mostly done with formula milk, whichlacks immunoreactive cells and immunoglobulins compared to breast milk. It is difficult to control the concentration and osmotic pressure during the preparation process, which increases the burden on the gastric mucosa of newborns to a certain extent and makes it easier for bacteriato invade and damage the intestinal wall [21].

In addition, this study also suggests that prenatal use of dexamethasone, breastfeeding, probiotics, use of PS, and platelet distribution width>11.8 fL are protective factors for NEC.Reasonableuse of dexamethasone before delivery can reduce the incidence of NEC, which may be related to the hormone promoting intestinal development [16], consistent with the study by Wang et al [27].Breast milk contains a wide range of microbiota and a variety of biologically active components that contribute to the development of the infant’s mucosal immune system, making itthe ideal natural food for infants [14,28].A study has shown that the use of probiotics is a method to regulate the gut microbiota of premature infants[29], which may be related to probiotics increasing mucus secretion, reducing mucosal permeability, enhancing mucosal barrier integrity, inhibiting intestinal bacterial translocation, regulating the number of T cells in the ileum, and promoting the secretion of immunoglobulin A in the intestinal mucosa [30].At present, there is limited research on the role and mechanism of PS and platelet distribution width in preventing NEC, and only one article [11] in this study reported that PS and platelet distribution width are protective factors of NEC, and their conclusions need further investigation.

### 4.3 The application prospects of NEC risk prediction models based on machine learning algorithms are promising

Machine learning is a branch of artificial intelligence that learns sample data through algorithms to accomplish specific tasks.With the development of artificial intelligence, machine learning theories such as random forests, artificial neural networks, multi-layer neural networks, and support vector machines have been widely applied in the research of disease diagnosis and prediction models, and have achieved ideal results [31].On the basis of the continuous development and improvement of medical information technology, the prevention and management of NEC is about to enter the era of big data. However, in the face of complex and massive data and the diversity of NEC risk prediction models, the application of traditional data analysis methods may not meet clinical needs.Machine learning can combine massive data with clinical information to develop NEC risk prediction models, achieving automatic, intelligent, and real-time analysis [19].In this study, a total of 2 articles [10,17] were risk prediction models developed based on machine learning algorithms, and the prediction results were still satisfactory.A systematic review by McAdams et al[32] showed that machine learning algorithms have shown outstanding performance in predicting neonatal diseases.However, machine learning still faces great challenges in the study of NEC risk prediction modeling, such as data generalizability, sample size, data sources, lack of model accuracy, poor fit, and ethics [31].It is suggested that future research should focus more on multicenter, large-scale data, and prospective research, continuously improving relevant laws and regulations to better address ethical issues.

## 5. Conclusion

This study found through a scope review method that the predictive performance of NEC risk prediction models is relatively good, but there is often a lack of external validation, resulting in an overall high risk of bias in China. Future clinical healthcare professionals should focus on high-risk factors for NEC, draw on the PROBAST evaluation criteria, develop and validate existing models, or use machine learning algorithms to construct models with good operability and predictive performance.

## Data Availability

All relevant data are within the manuscript and its Supporting Information files.

